# Clinical Characteristics on 25 Discharged Patients with COVID-19 Virus Returning

**DOI:** 10.1101/2020.03.06.20031377

**Authors:** Jing Yuan, Shanglong Kou, Yanhua Liang, JianFeng Zeng, Yanchao Pan, Lei Liu

**Affiliations:** Diagnosis and Treatment of Infectious Diseases Research Laboratory, Shenzhen Third People’s Hospital, Shenzhen 518112, China

## Abstract

Here we report the clinical features of 25 discharged patients with COVID-19 recovery. Our analysis indicated that there was a significant inverse correlation existed between serum D-Dimer level and the duration of antiviral treatment, while lymphocyte concentration significantly positively correlated with the duration of virus reversal.

## Introduction

Since Dec 8^th^ 2019, many cases has been reported by investigators who described the clinical characteristics of hospitalized patients with COVID-19 infection^1-3^. We noticed the recent report^4^ from Lan et al. that 4 medical staff were still virus carriers after recovery from COVID-19 infection. Here we gave a report on 25 discharged patients with their medical record review and further analysis.

## Methods

All these 25 patients with COVID-19 infection were once hospitalized from Jan 23^th^ 2020 to Feb 21^th^ 2020. They all met the following criteria of hospital discharge in China: (1) more than 3 days of normal; (2) significant reduction of respiratory symptoms; (3) substantial improvement over conventional chest radiography detection; (4) consecutively twice negative results of RT-PCR testing with an interval for at least 1 day. Considering the risk of reinfection, they were all self-segregating at home for further observation. Real-time RT-PCR detection were performed as described previously^5^. Other demographic, clinical, radiologic and laboratory findings were extracted from the electronic medical records of the patients. The study was approved by Shenzhen Third People’s Hospital Ethics Committee and the informed consent was waived.

## Results

25 discharged patients with COVID-19 infection were back hospitalized because of the virus mRNA recurrence. The median age of these 17 females and 8 males was 28 years (16.25∼42), including 6 children under 12 years old. 21 of them had no comorbidity and 22 patients had the history of residing in Hubei province. Besides, 24 of them were once non-severe patient with the common symptoms of fever (17/25) and cough (14/25) at onset. Overall, with 14 (13-18.5) days of the hospital stay, as well as 13 (10.5-16.5) days of antivirus treatment, the patients all discharged in 2 (1-3.8) days after two consecutive negative results on virus mRNA RT-PCR detection, as well as improvements on chest computed tomography (CT) evidence.

With 14/25 of cloacal swab samples and rest of Nasal Swabs or oropharynx swab samples testing, these patients presented the repeated positive COVID-19 mRNA within 3 (2-7) days after the hospital. Notably, the median time from their last negative result to turning positive was 6 (4-10) days. These patients were then hospitalized again and continued the quarantine protocol. All these patients were asymptomatic and chest CT scanning indicated that 12 of them even showed improvement while rest of them represented with no obvious change compared with previous images. With a few days of prophylactic intervention with Chinese herbal medicine, the RT-PCR results of virus mRNA detection were all turning to negative in both nasopharyngeal swab and cloacal swab samples.

Furthermore, correlation analysis indicated that there was a significant inverse correlation existed between serum D-Dimer level and the duration of antiviral treatment (r=-0.637, *p*=0.002), while lymphocyte concentration significantly positively correlated (r=0.52, *p*=0.008) with the duration of virus reversal. These implied that the imperfect antivirus therapy probably was responsible for the recurrence of COVID-19 virus.

## Discussion

These 25 patients with COVID-19 infection all met the criteria for hospital release from quarantine, while the RT-PCR testing then conversed to positive without symptoms after 2 to 13 days. These asymptomatic carriers brought more challenges to the manager and control of COVID-19 epidemic in China and any other affected countries. We further got a glance at the factors associated with COVID-19 clearance. Due to the lack of antiviral drugs with confirmed efficiency on COVID-19, it is possible that some immunological parameters could be used as additional indicators to properly assess the virus activity and the needs for prolonged quarantine in asymptomatic patients. Further case-control study and cohort study will be needed to pursue the indication role on that.

## Data Availability

Anyone who wishes to share, reuse, remix, or adapt this material please send email to the corresponding author.

## Author Contributions

Dr. Yanchao Pan and Lei Liu had full access to all of the data in the study and take responsibility for the integrity of the data and the accuracy of the data analysis. Acquisition, analysis, or interpretation of data: Jing Yuan, Yanhua Liang, Jianfeng Zeng. Concept and design: Jing Yuan, Lei Liu. Drafting of the manuscript: Yanchao Pan, Shanglong Kou. Statistical analysis: Yanchao Pan, Shanglong Kou.

---

**Table 1.** Baseline Characteristics of 25 Discharged Patients with COVID-19 return.

| Age Range |  | Last hospital stay | antivirus treatment durations | Time from negative PCR test to discharge | Time from positive again to last negative | Time from positive again to last discharge | Time from positive again to second hospitalization |
| --- | --- | --- | --- | --- | --- | --- | --- |
| ≤20 | Average Days | 15 | 13.33 | 3.67 | 8.33 | 4.67 | 1.33 |
|  | SD | 2.828 | 3.933 | 2.503 | 5.61 | 4.227 | 1.506 |
|  | N | 6 | 6 | 6 | 6 | 6 | 6 |
| 20-40 | Average Days | 15.23 | 13.46 | 2.17 | 6.77 | 5.92 | 1.85 |
|  | SD | 3.345 | 4.313 | 1.528 | 3.395 | 4.716 | 2.115 |
|  | N | 13 | 13 | 12 | 13 | 13 | 13 |
| 41-60 | Average Days | 16 | 13.5 | 2.83 | 7.5 | 4.67 | 2.67 |
|  | SD | 5.831 | 4.461 | 1.722 | 3.209 | 2.944 | 2.066 |
|  | N | 6 | 6 | 6 | 6 | 6 | 6 |
| Total | Average Days | 15.36 | 13.44 | 2.71 | 7.32 | 5.32 | 1.92 |
|  | SD | 3.807 | 4.083 | 1.876 | 3.859 | 4.13 | 1.956 |
|  | N | 25 | 25 | 24 | 25 | 25 | 25 |

Data are given as mean ± standard deviation (SD).

**Figure 1.**
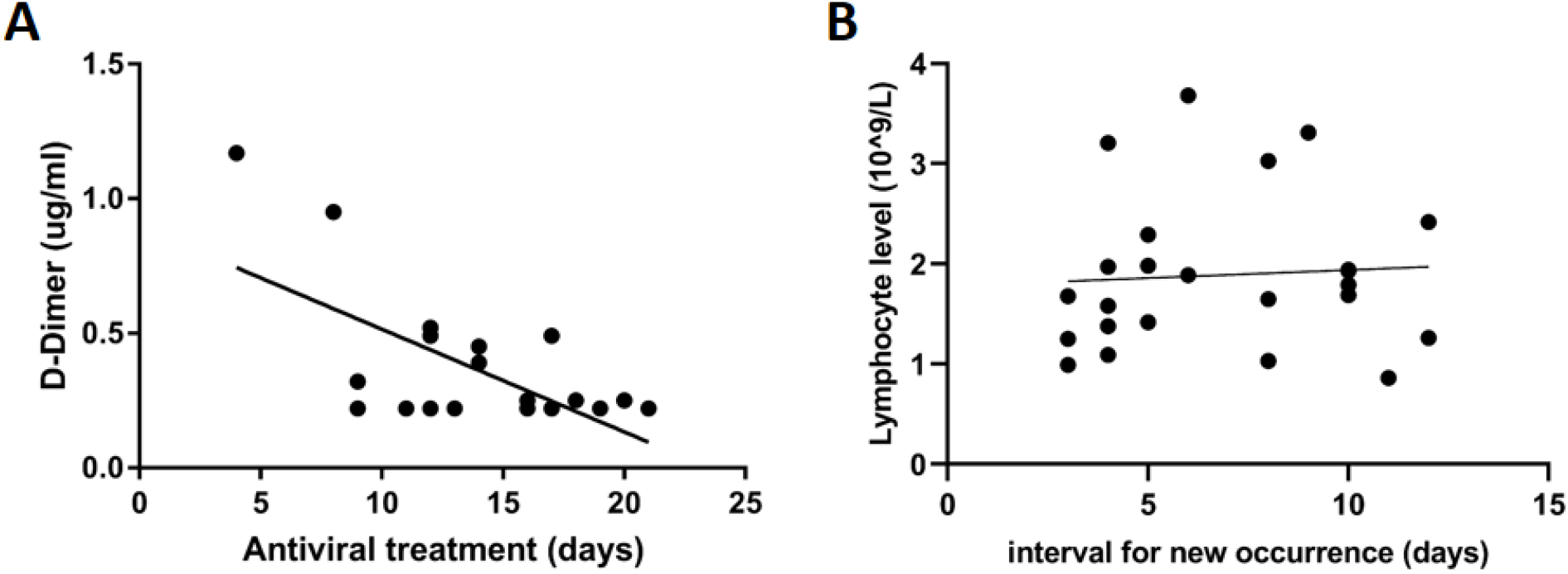
Correlation analysis for lymphocyte count and duration of virus reversal, as well as serum D-Dimer level and duration of antiviral treatment. Correlation analysis on serum D-Dimer level and the duration of antiviral treatment (r=-0.637, *p*=0.002) (A), as well as correlation between lymphocyte concentration significantly and the duration of virus reversal (r=0.52, *p*=0.008) (B).

